# Cross-Sectional Measures of Periodontal Severity: Distortion from Severity-Dependent Tooth Loss

**DOI:** 10.64898/2026.05.27.26354277

**Authors:** Kym M. McCormick, Najith Amarasena, Gina Guzzo, Sonia Nath, Lisa Jamieson

## Abstract

**Aim:** Cross-sectional summaries of periodontitis based on clinical attachment loss (CAL) are, by definition, conditioned on surviving teeth. Because the most severely affected teeth are more likely to have been lost, these measures may underestimate cumulative disease burden and show an artificial flattening (attenuation) of severity with age. We hypothesised that measures more sensitive to severe attachment loss would show greater attenuation at older ages than measures defined across a broader range of sites.

**Materials and Methods:** Using nationally representative data from adults aged ≥30 years in NHANES 2009–2014, we examined age-specific trajectories across multiple continuous measures of periodontal severity and assessed whether divergence between measures followed the pattern predicted under severity-dependent tooth loss.

**Results:** The proportion of observable sites declined from 93% at ages 30–34 to 68% at ≥80 years, establishing the structural basis for the divergence observed across severity measures. All severity measures showed nonlinear attenuation with age, with distortion increasing with severity threshold. Higher-threshold measures exhibited the greatest attenuation, while lower-threshold measures showed more stable trajectories.

**Conclusions:** Cross-sectional summaries of periodontitis reflect disease among surviving teeth rather than cumulative damage across teeth originally at risk. Attenuation at older ages is consistent with depletion of the most severely affected teeth rather than biological slowing. Distortion varies by measure, with higher-threshold and mean-based indices most affected, whereas the CAL ≥3 mm threshold provides a more stable basis for age comparisons.

## Introduction

Periodontitis is a chronic inflammatory disease characterised by progressive and largely irreversible destruction of the supporting structures of the teeth, ultimately resulting in tooth loss (Pihlstrom et al. 2005; Kinane et al. 2017; Kassebaum et al. 2014). Because this destruction accumulates over decades, the burden of disease at any given time reflects the cumulative history of tissue breakdown across the dentition rather than current inflammatory activity alone. Its quantification therefore depends critically on how cumulative damage is measured. Clinical attachment loss (CAL) at retained teeth is the standard index of periodontal destruction and provides a continuous representation of disease severity across individuals and populations (Page and Eke 2007; Dye et al. 2019; Du et al. 2023). Unlike threshold-based case definitions, which summarise disease status relative to fixed cut-points, continuous measures of CAL preserve information on the distribution and gradient of periodontal breakdown within the population.

The usefulness of continuous measures of CAL for representing cumulative periodontal destruction depends on an implicit assumption: that the teeth available for examination adequately reflect the full history of disease across the dentition. In cross-sectional studies, however, CAL can only be assessed at teeth present at the time of examination. As periodontitis progresses, teeth with the greatest attachment loss are more likely to be lost, whether through disease progression or clinical intervention (Ong 1998; Müller et al. 2007; Haworth et al. 2018; Serino et al. 2001). Once lost, the attachment loss they carried is no longer observable. Consequently, periodontal assessments—particularly in older adults—are conducted on a dentition that has been selectively depleted of its most severely affected teeth. This process removes the most severely affected teeth from observation, leaving a dentition that under-represents the true extent of periodontal destruction.

This process has predictable consequences for how periodontal severity is observed. As the most severely affected teeth are lost over time, the remaining dentition can appear less severely affected than the cumulative burden of destruction would suggest. Apparent attenuation or plateauing of CAL at older ages may therefore reflect the selective loss of high-burden teeth rather than any true slowing of disease progression. This effect will be more pronounced for measures that depend on the most severely affected sites (e.g., percentage of sites with CAL ≥6 mm), which are directly implicated in tooth loss, than for measures defined across a broader range of sites (e.g., CAL ≥3 mm), which are less sensitive to the loss of extreme values (McCormick 2026). As a result, different continuous measures applied to the same population may show diverging age-related patterns, with the extent of divergence reflecting each measure’s sensitivity to severe disease.

These considerations have implications for how periodontal severity is interpreted in both research and epidemiological contexts. Previous work has noted that tooth loss may complicate the interpretation of periodontal measures in older populations and may contribute to apparent attenuation of associations in epidemiological studies (Hujoel et al. 2000; Dietrich et al. 2008). However, the extent to which severity-linked tooth loss distorts commonly used continuous indices, and whether it produces systematic divergence between indices with differing sensitivity to severe attachment loss, has not been well quantified.

Using nationally representative data from NHANES 2009–2014, we examined age-specific patterns across multiple continuous measures of periodontal severity and assessed whether divergence between these measures follows the pattern expected under severity-dependent tooth loss. We hypothesised that measures more sensitive to severe attachment loss would show greater attenuation at older ages than measures defined across a broader range of sites.

## Methods

### Data Source and Study Population

We analysed data from the 2009–2014 National Health and Nutrition Examination Survey (NHANES), combining three 2-year cycles (2009–2010: n = 3,752; 2011–2012: n = 3,338; 2013–2014: n = 3,624), a nationally representative, multistage probability sample of the U.S. civilian noninstitutionalized population (Dye et al. 2019; Dye et al. 2014). These cycles were selected as they represent the most recent NHANES data that include full-mouth protocols with six sites per tooth (mesiobuccal, buccal, distobuccal, mesiolingual, lingual, distolingual), excluding third molars. Subsequent cycles employed a partial-mouth design that limits comparability.

Analyses were restricted to adults aged ≥30 years who completed the periodontal examination. Edentulous participants were not included. In the pooled analytic sample, the weighted mean age was 50.8 years (SE 0.24), and 51.2% (SE 0.51) were female. Six-year examination weights were constructed by combining three 2-year cycles according to National Center for Health Statistics guidelines.

NHANES protocols were approved by the National Center for Health Statistics Research Ethics Review Board, and all participants provided written informed consent. The present analyses used publicly available, de-identified data and did not require additional ethics review.

## Measures

### Structural observability

Structural observability was defined at the tooth/site level. For each participant, we calculated the number of natural teeth present and the proportion of eligible periodontal sites with recorded CAL measurements as measures of structural observability. Eligible sites were defined as the 168 sites comprising a full dentition (28 teeth × 6 sites). This reflects both tooth loss and any site-level missingness in the examination protocol.

### Periodontal severity

Full-mouth severity summaries were computed using available CAL measurements at observed sites only (i.e., excluding missing teeth and sites without recorded CAL). Outcomes included the percentage of sites with CAL ≥3 mm, ≥5 mm, and ≥6 mm, and mean CAL (mm), consistent with established surveillance definitions (Page and Eke 2007; Eke et al. 2012; Du et al. 2023). No attempt was made to impute attachment levels for missing teeth; summaries therefore reflect severity among retained structures rather than the full lifetime burden of disease.

## Statistical analysis

Age-specific weighted means and 95% confidence intervals were estimated using survey-weighted procedures accounting for NHANES strata, primary sampling units, and examination weights.

To assess departures from linear age–severity relationships, we fitted survey-weighted regression models for each severity measure with age entered as an ordinal index (1–11) corresponding to ordered age groups (30–34 through ≥80 years). For each outcome, both linear and quadratic specifications were estimated. Nonlinearity was formally tested using Rao–Scott likelihood ratio tests comparing quadratic to linear models, with survey design– adjusted degrees of freedom (*df* =46).

In quadratic models, the coefficient on the squared age term (β_2_) was used to characterise curvature. A negative β_2_ indicates attenuation of the age–severity trajectory at older ages. For descriptive interpretation, implied peak ages were calculated as -β_1_/(2β_2_).

To examine the relationship between mean clinical attachment loss (CAL) and the percentage of sites with CAL ≥3 mm across levels of structural retention, participants were stratified by tooth count (<20, 20–24, ≥25), reflecting clinically meaningful thresholds of dentition retention, with <20 teeth commonly used to define a non-functional dentition (WHO, 2013). Within each stratum, the association between mean CAL and the ≥3 mm extent measure was modelled using a survey-weighted quasibinomial generalised linear model with mean CAL entered as a continuous predictor, appropriate for the bounded, proportional outcome.

All analyses were conducted in R (version 4.5.1; R Core Team, Vienna, Austria) using the survey package (Lumley 2004). Model-based curves in Figure 2 were derived from these quasibinomial models and plotted using ggplot2 (Wickham 2016).

## Results

Results are presented to describe (i) how many teeth and sites remain observable, (ii) how periodontal severity changes with age, and (iii) how different severity measures relate to each other at different levels of tooth retention.

### Descriptive patterns

Table 1 shows the characteristics of the study population by age group. The proportion of observable sites decreased steadily with age, from 93.2% in adults aged 30–34 years to 67.9% in those aged ≥80 years. This decline reflects the progressive loss of teeth (mean: 26.5 to 19.4), and therefore a reduction in the structures available for measurement.

**Table 1.**
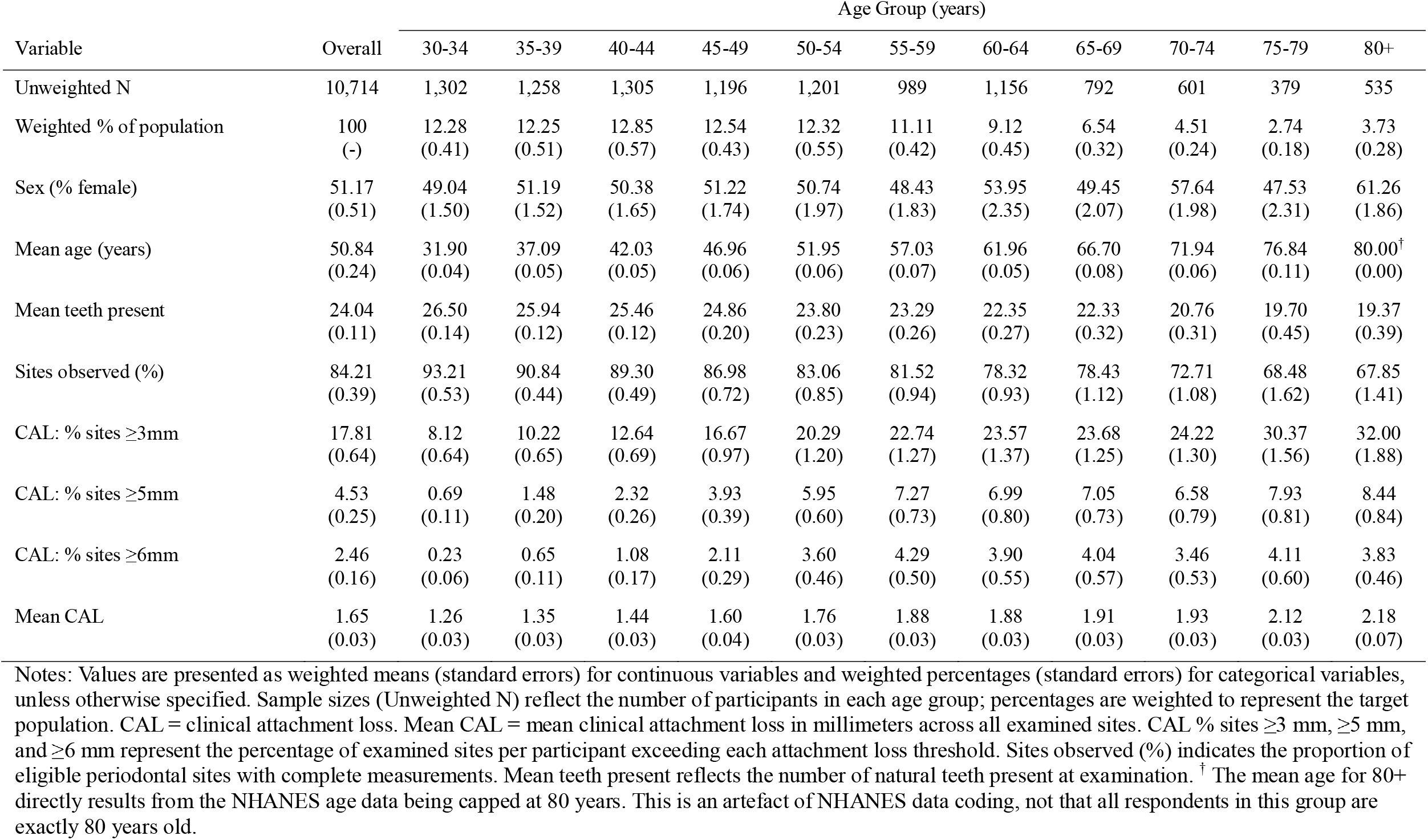
Weighted characteristics of the study population by age group (years).

| Variable | Overall | Age Group (years) |  |  |  |  |  |  |  |  |  |  |
| --- | --- | --- | --- | --- | --- | --- | --- | --- | --- | --- | --- | --- |
|  |  | 30-34 | 35-39 | 40-44 | 45-49 | 50-54 | 55-59 | 60-64 | 65-69 | 70-74 | 75-79 | 80+ |
| Unweighted N | 10,714 | 1,302 | 1,258 | 1,305 | 1,196 | 1,201 | 989 | 1,156 | 792 | 601 | 379 | 535 |
| Weighted % of population | 100<br>(-) | 12.28<br>(0.41) | 12.25<br>(0.51) | 12.85<br>(0.57) | 12.54<br>(0.43) | 12.32<br>(0.55) | 11.11<br>(0.42) | 9.12<br>(0.45) | 6.54<br>(0.32) | 4.51<br>(0.24) | 2.74<br>(0.18) | 3.73<br>(0.28) |
| Sex (% female) | 51.17<br>(0.51) | 49.04<br>(1.50) | 51.19<br>(1.52) | 50.38<br>(1.65) | 51.22<br>(1.74) | 50.74<br>(1.97) | 48.43<br>(1.83) | 53.95<br>(2.35) | 49.45<br>(2.07) | 57.64<br>(1.98) | 47.53<br>(2.31) | 61.26<br>(1.86) |
| Mean age (years) | 50.84<br>(0.24) | 31.90<br>(0.04) | 37.09<br>(0.05) | 42.03<br>(0.05) | 46.96<br>(0.06) | 51.95<br>(0.06) | 57.03<br>(0.07) | 61.96<br>(0.05) | 66.70<br>(0.08) | 71.94<br>(0.06) | 76.84<br>(0.11) | 80.00<sup>†</sup><br>(0.00) |
| Mean teeth present | 24.04<br>(0.11) | 26.50<br>(0.14) | 25.94<br>(0.12) | 25.46<br>(0.12) | 24.86<br>(0.20) | 23.80<br>(0.23) | 23.29<br>(0.26) | 22.35<br>(0.27) | 22.33<br>(0.32) | 20.76<br>(0.31) | 19.70<br>(0.45) | 19.37<br>(0.39) |
| Sites observed (%) | 84.21<br>(0.39) | 93.21<br>(0.53) | 90.84<br>(0.44) | 89.30<br>(0.49) | 86.98<br>(0.72) | 83.06<br>(0.85) | 81.52<br>(0.94) | 78.32<br>(0.93) | 78.43<br>(1.12) | 72.71<br>(1.08) | 68.48<br>(1.62) | 67.85<br>(1.41) |
| CAL: % sites ≥3mm | 17.81<br>(0.64) | 8.12<br>(0.64) | 10.22<br>(0.65) | 12.64<br>(0.69) | 16.67<br>(0.97) | 20.29<br>(1.20) | 22.74<br>(1.27) | 23.57<br>(1.37) | 23.68<br>(1.25) | 24.22<br>(1.30) | 30.37<br>(1.56) | 32.00<br>(1.88) |
| CAL: % sites ≥5mm | 4.53<br>(0.25) | 0.69<br>(0.11) | 1.48<br>(0.20) | 2.32<br>(0.26) | 3.93<br>(0.39) | 5.95<br>(0.60) | 7.27<br>(0.73) | 6.99<br>(0.80) | 7.05<br>(0.73) | 6.58<br>(0.79) | 7.93<br>(0.81) | 8.44<br>(0.84) |
| CAL: % sites ≥6mm | 2.46<br>(0.16) | 0.23<br>(0.06) | 0.65<br>(0.11) | 1.08<br>(0.17) | 2.11<br>(0.29) | 3.60<br>(0.46) | 4.29<br>(0.50) | 3.90<br>(0.55) | 4.04<br>(0.57) | 3.46<br>(0.53) | 4.11<br>(0.60) | 3.83<br>(0.46) |
| Mean CAL | 1.65<br>(0.03) | 1.26<br>(0.03) | 1.35<br>(0.03) | 1.44<br>(0.03) | 1.60<br>(0.04) | 1.76<br>(0.03) | 1.88<br>(0.03) | 1.88<br>(0.03) | 1.91<br>(0.03) | 1.93<br>(0.03) | 2.12<br>(0.03) | 2.18<br>(0.07) |

Notes: Values are presented as weighted means (standard errors) for continuous variables and weighted percentages (standard errors) for categorical variables, unless otherwise specified. Sample sizes (Unweighted N) reflect the number of participants in each age group; percentages are weighted to represent the target population. CAL = clinical attachment loss. Mean CAL = mean clinical attachment loss in millimeters across all examined sites. CAL % sites ≥3 mm, ≥5 mm, and ≥6 mm represent the percentage of examined sites per participant exceeding each attachment loss threshold. Sites observed (%) indicates the proportion of eligible periodontal sites with complete measurements. Mean teeth present reflects the number of natural teeth present at examination. <sup>†</sup> The mean age for 80+ directly results from the NHANES age data being capped at 80 years. This is an artefact of NHANES data coding, not that all respondents in this group are exactly 80 years old.

As structural loss occurs, patterns of periodontal severity change. The percentage of sites with CAL ≥3 mm increased steadily across age groups, consistent with ongoing accumulation of attachment loss. In contrast, measures based on higher thresholds showed a different pattern. The percentage of sites with CAL ≥6 mm increased through midlife, then declined modestly in the oldest groups. Mean CAL also increased with age, but the rate of increase slowed at older ages (Figure 1).

**Figure 1.**
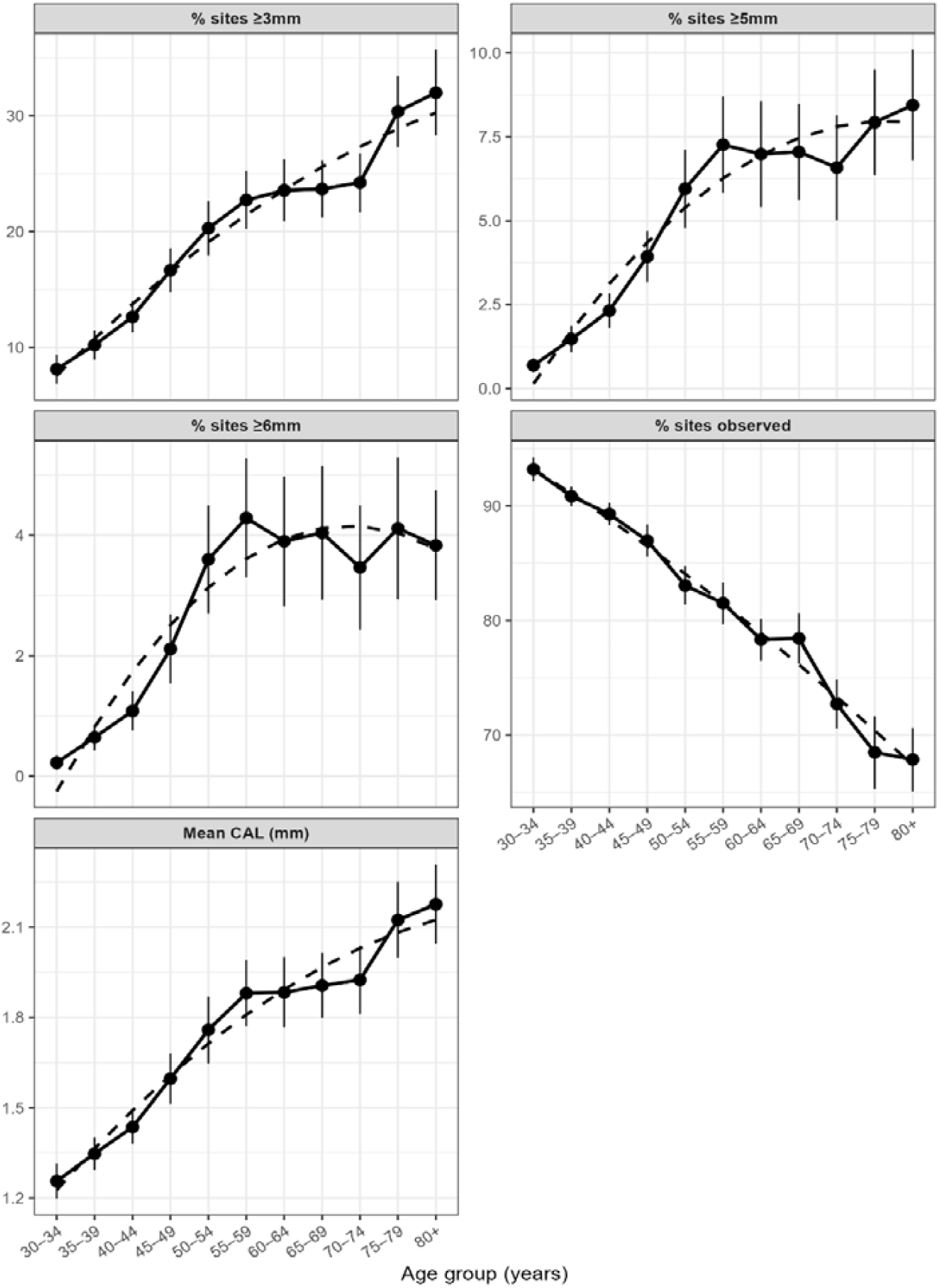
Observed age-specific periodontal measures with survey-weighted quadratic model estimates. Points represent weighted observed estimates by age group (30–34 to ≥80 years), with vertical bars indicating standard errors. Dashed lines represent survey-weighted quadratic model estimates. Panels display percentage of sites with CAL ≥3 mm, ≥5 mm, and ≥6 mm; percentage of sites observed; and mean clinical attachment loss (CAL, mm). Across age groups, observed and model estimates show similar monotonic increases in periodontal severity and a decline in the proportion of sites observed. Nonlinear attenuation at older ages, most pronounced for higher severity thresholds, is consistent with progressive severity-dependent loss of the most affected teeth from the observable dentition rather than biological deceleration of disease accumulation.

These patterns suggest that different measures of severity do not behave equivalently across age, particularly as the number of remaining teeth declines.

### Nonlinearity

Statistical tests showed that a curved (quadratic) model fit the data better than a straight-line model for all measures (Table 2). In each case, the curve bent downward at older ages, indicating attenuation, or a slowing and in some cases reversal of the observed increase in severity.

**Table 2.**
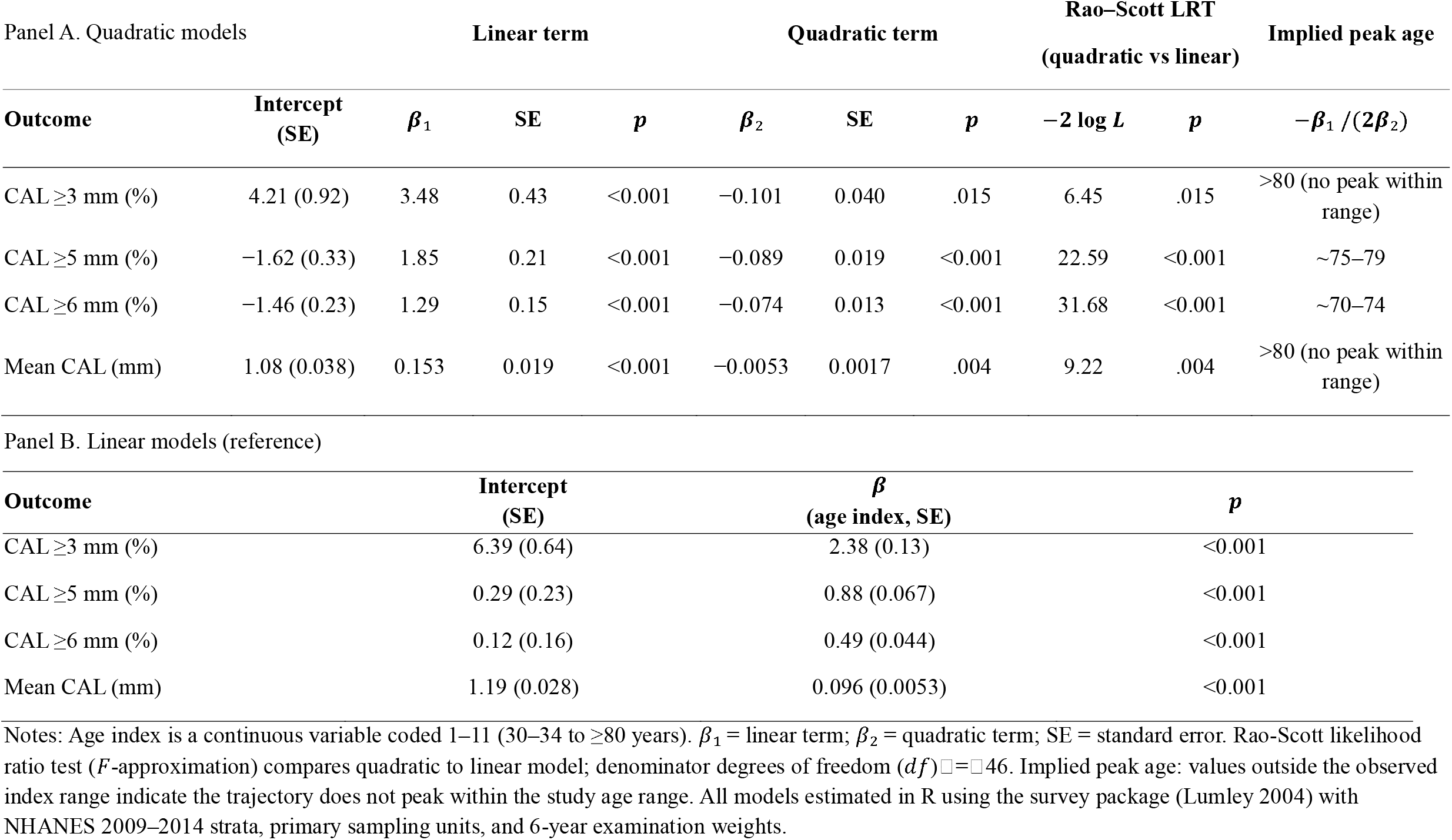
Survey-weighted regression model results for age–periodontal severity associations.

The extent of this curvature depended on the severity threshold. For the ≥6 mm measure, values increased rapidly through midlife, peaked in the early 70s, and then declined. This indicates that the most severe disease becomes progressively underrepresented at older ages. The ≥5 mm measure showed a similar but less pronounced pattern, with a peak near the oldest age groups.

These peak points are clinically informative. They mark the stage at which further cumulative attachment loss is no longer captured in cross-sectional summaries, because the teeth most affected by severe disease have already been lost.

In contrast, the ≥3 mm and mean CAL measures continued to increase across all age groups, although more slowly at older ages, and did not show a clear peak within the observed range. This difference across measures forms a consistent pattern: the more a measure depends on severely affected sites, the more it is affected by their loss.

Because cumulative attachment loss cannot decrease over time, these patterns are consistent with selective removal of the most severely affected teeth from the observable dentition in later life, when periodontal tooth loss becomes more common (Müller et al. 2007).

### Relationship between mean CAL and extent of attachment loss ≥3 mm

Figure 2 shows how mean CAL relates to the percentage of sites with CAL ≥3 mm, grouped by number of teeth present. Across all groups, a common pattern was observed. At low mean CAL values, the proportion of affected sites increased gradually. At moderate levels, the increase became steeper. At higher values, the curve levelled off, indicating that most sites were already affected.

**Figure 2.**
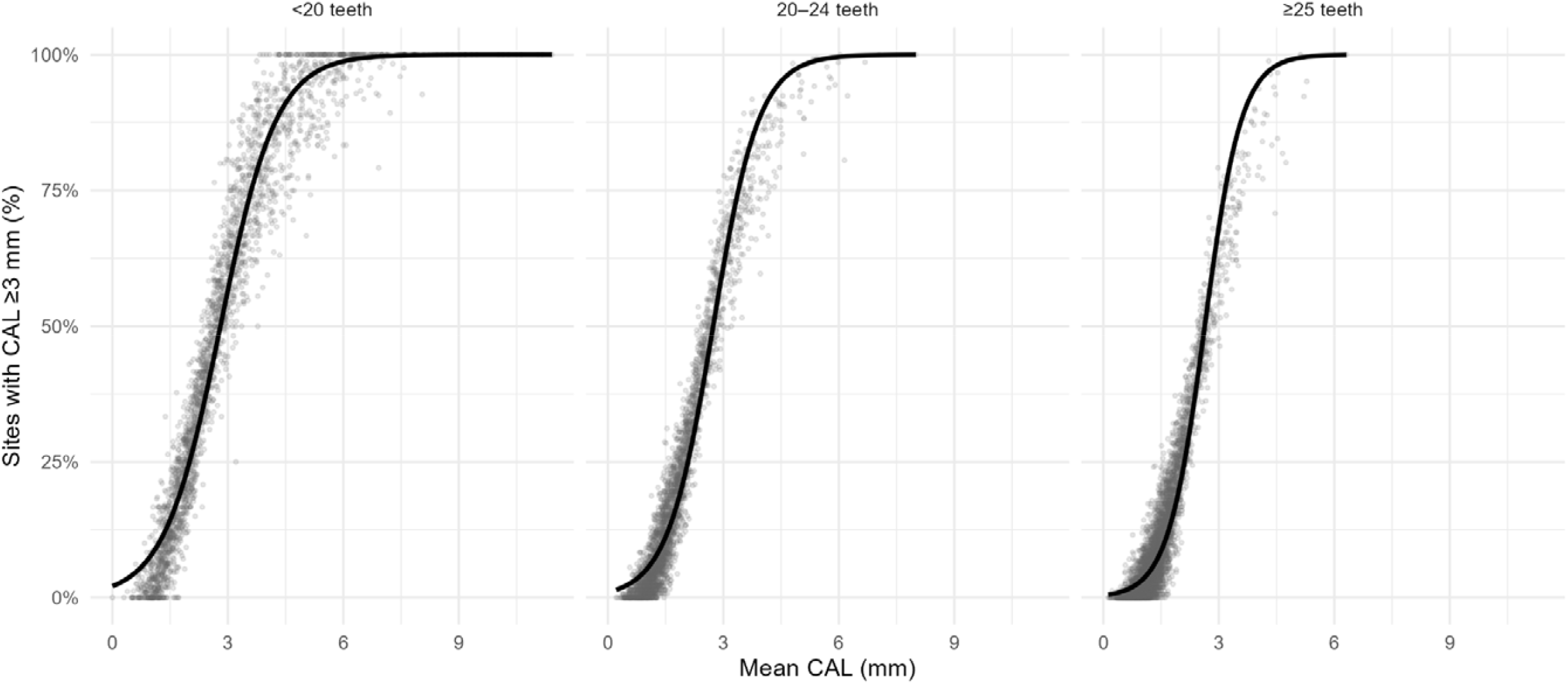
Relationship between mean clinical attachment loss (CAL) and percentage of sites with CAL ≥3 mm, stratified by tooth count. Tooth count strata reflect clinically meaningful levels of dentition retention, with <20 teeth commonly used to define a non-functional dentition (World Health Organization, 2013). Each point represents one participant from NHANES 2009–2014. Lines show quasibinomial generalised linear model fits of the relationship between mean CAL and the proportion of sites with CAL ≥3 mm within each stratum. In individuals with ≥25 teeth, the two measures track closely across the observed range. As tooth count decreases, this relationship becomes non-invariant: in individuals with fewer than 20 teeth, the curve plateaus at lower mean CAL values, with further increases in mean CAL associated with little additional increase in affected sites. This pattern is consistent with compression of the observable disease distribution under declining structural retention, where high-burden teeth are no longer represented in the extent measure denominator. The divergence across strata indicates that mean CAL and threshold-based extent measures are not interchangeable indices of periodontal burden, and that their relationship depends on the level of structural retention.

Although this general shape was consistent, its behaviour differed by tooth count. Among individuals with ≥25 teeth, the curve rose steeply across the full range, indicating that the extent measure remained sensitive to variation in mean CAL. In those with 20–24 teeth, the curve began to flatten at higher mean CAL values, suggesting a loss of sensitivity at higher levels of disease.

The most pronounced effect was seen in individuals with fewer than 20 teeth. In this group, the proportion of sites with CAL ≥3 mm approached an upper limit, even as mean CAL continued to increase. This indicates that the extent measure becomes compressed when fewer teeth remain, limiting its ability to distinguish between higher levels of disease.

In practical terms, the extent measure is most informative at lower levels of disease, when fewer sites are affected. As disease becomes more widespread and tooth loss progresses, its ability to differentiate severity diminishes. As a result, the relationship between mean CAL and extent depends on how many teeth are present, and cannot be interpreted independently of structural loss.

## Discussion

We predicted that measures more sensitive to severe attachment loss would show greater attenuation at older ages than measures defined across a broader range of sites. This prediction was supported. Importantly, attenuation did not occur uniformly across measures, but followed a predictable hierarchy determined by each measure’s sensitivity to severe disease. Cross-sectional severity trajectories showed clear attenuation and nonlinearity with age, with distortion most pronounced for higher severity thresholds.

Similar patterns have previously been interpreted as reflecting age-dependent changes in disease distribution or biological slowing of progression (Billings et al. 2018; Eke et al. 2015). The present findings suggest a different explanation. The observed hierarchy follows directly from a structural selection process operating at the level of the dentition, whereby the most severely affected teeth are preferentially lost over time. As a result, cross-sectional measures are necessarily conditioned on the subset of teeth that remain observable at examination.

By construction, these measures describe periodontal burden among retained teeth rather than the full extent of cumulative destruction, such that attenuation at older ages reflects progressive depletion of high-burden sites rather than a slowing of disease.

Observed patterns of threshold-dependent attenuation reflect how strongly each severity measure couples with tooth loss. Sites meeting the ≥6 mm threshold represent the most severely affected portion of the dentition and are therefore most likely to be lost, whether through disease progression or clinical extraction (Tonetti et al. 2018). As these sites are preferentially removed from the observable dentition, the ≥6 mm measure becomes increasingly disconnected from the underlying burden at older ages, producing the marked plateau observed in the NHANES data. In contrast, the ≥3 mm threshold captures a broader range of affected sites, including those on teeth not yet at risk of extraction. As a result, the denominator is less truncated and the measure retains a closer relationship to cumulative disease, although it remains conditioned on surviving teeth and continues to underestimate true lifetime burden as tooth loss progresses.

Mean clinical attachment loss (CAL) is often assumed to provide a straightforward summary of periodontal disease severity, but its behaviour under tooth loss is less intuitive than threshold-based measures. Mean CAL, however, presents a different and less visible version of the same problem. Like extent measures, it is computed over surviving teeth only, and its denominator shrinks non-randomly as the most severely affected teeth are lost. Unlike extent measures, which are bounded and show visible attenuation when severely affected sites are removed, mean CAL has no natural upper limit. Two processes therefore operate simultaneously in opposite directions: ongoing attachment loss in surviving teeth pushes the estimate upward, while progressive loss of high-burden teeth removes the observations that would most increase it. These forces can partially offset one another, producing a smooth, apparently well-behaved trajectory that gives little indication that structural truncation is operating. The measure continues to increase, but at a rate reflecting the net effect of accumulation and depletion rather than accumulation alone. As a result, identical mean CAL values may reflect fundamentally different underlying disease histories, such that the measure does not uniquely represent cumulative periodontal destruction.

This cancellation is supported by the observed relationship between mean CAL and the ≥3 mm extent measure stratified by tooth count (Figure 2). As tooth count decreases, the mapping between the two measures becomes non-invariant, with higher mean CAL values corresponding to progressively lower percentages of sites with CAL ≥3 mm. This pattern is consistent with compression of the observable disease distribution as structural retention declines.

### Implications for exposure measurement

Research linking periodontitis to systemic conditions is premised on cumulative inflammatory burden as the pathogenic mechanism, rather than current clinical status (Herrera et al. 2023). Measures used as exposures in this context therefore aim to capture accumulated damage. Cross-sectional CAL measures are commonly used for this purpose, but in older populations what is observed at examination reflects severity among teeth that remain, not the total damage accrued over a lifetime. Individuals with the greatest cumulative exposure may have already lost the teeth that would most clearly reflect that history. This has implications for effect estimation: if periodontal exposure is underestimated most severely among those with the highest true burden, associations with systemic outcomes may be attenuated toward the null. This interpretation is consistent with evidence that periodontitis– mortality associations are often modest and that cause-specific effects are frequently non-significant in older or high-comorbidity populations (Xu and Lu 2011; Chen et al. 2024), a pattern that may arise, at least in part, from severity-dependent structural truncation rather than solely from age-related changes in the underlying biological relationship.

Mean CAL does not directly estimate cumulative attachment loss across the original dentition. Rather, it reflects mean attachment loss among teeth that survived long enough to be measured; a quantity whose relationship to true cumulative burden depends on the extent and severity-dependence of tooth loss. This distinction is especially consequential for disparities research. Groups that experience earlier or more severe tooth loss — including those with lower socioeconomic position, limited access to dental care, or higher systemic disease burden — may present with a more severely depleted tooth set at any given age, not necessarily because their underlying disease experience is less severe, but because the most severely affected teeth have already been lost (McCormick et al. 2025; Thomson 2012; Müller et al. 2007). Cross-sectional comparisons between such groups may therefore underestimate true differences in cumulative periodontal exposure, potentially biasing comparisons against those already most disadvantaged.

### Limitations and Future Directions

Several limitations warrant consideration. Cross-sectional data cannot recover counterfactual cumulative burden without additional assumptions, and the present analyses are descriptive rather than causal. We show that the observed patterns are consistent with severity-dependent structural truncation but cannot directly observe the joint process linking cumulative severity and tooth loss within individuals. The structural selection mechanism is therefore inferred analytically, and its underlying parameters are not identifiable from cross-sectional data alone.

Differential mortality may further compound structural depletion at the individual level. Individuals with higher periodontal burden are more likely to experience both early tooth loss and earlier mortality (Xu and Lu 2011; Kassebaum et al. 2014). As a result, older age groups in cross-sectional data are selected both for having retained more teeth and for having survived. These processes remove high-burden individuals and high-burden teeth from the observable set and cannot be disentangled without linked longitudinal and mortality data. The estimates reported here therefore reflect the combined influence of both mechanisms.

Recovering cumulative burden across the full original dentition requires explicit assumptions about the joint distribution of severity and structural loss. One approach combines observed CAL in retained teeth with probabilistic contributions from missing teeth, informed by age-stratified data on periodontal extraction. A further refinement would incorporate intraoral spatial patterns of tooth-level vulnerability. Such approaches move inference from implicit survivor-conditioning toward explicit estimation of cumulative burden and may better capture the exposure hypothesised to drive systemic disease associations (Herrera et al. 2023).

These measures would not replace survivor-conditioned summaries, which remain appropriate for describing current clinical status among retained structures. Rather, distinguishing these quantities clarifies which scientific question is being addressed and reduces the risk that mechanically induced patterns are interpreted as biological phenomena.

## Conclusion

Cross-sectional measures of periodontal severity reflect disease among surviving teeth rather than cumulative burden. Severity-dependent tooth loss is sufficient to produce the attenuation in age–severity trajectories often attributed to biological deceleration. The extent of this distortion depends on measure choice, with higher-threshold and mean-based measures most affected. These effects should be considered when interpreting cross-sectional periodontal data, particularly in older populations.

## Data Availability

All data produced are available online at https://wwwn.cdc.gov/nchs/nhanes/default.aspx

https://wwwn.cdc.gov/nchs/nhanes/search/datapage.aspx?Component=Examination&Cycle=2009-2010

https://wwwn.cdc.gov/nchs/nhanes/search/datapage.aspx?Component=Examination&Cycle=2011-2012

https://wwwn.cdc.gov/nchs/nhanes/search/datapage.aspx?Component=Examination&Cycle=2013-2014

